# Nephroprotective effects of cilastatin in people at risk of acute kidney injury: A systematic review and meta-analysis

**DOI:** 10.1101/2024.03.06.24303823

**Authors:** Dilaram Acharya, Fanar Ghanim, Tyrone G. Harrison, Tayler Dawn Scory, Nusrat Shommu, Paul E. Ronksley, Meghan J. Elliott, David Collister, Neesh Pannu, Matthew T. James

## Abstract

**Rationale & Objective:** Cilastatin is an inhibitor of drug metabolism in the proximal tubule that demonstrates nephroprotective effects in animals. It has been used in humans in combination with the antibiotic imipenem to prevent imipenem’s degradation. This systematic review and meta-analysis evaluated the nephroprotective effects of cilastatin in humans.

**Study Design:** Systematic review and meta-analysis of observational (comparative effectiveness) studies or randomized clinical trials (RCTs)

**Setting & Study Populations:** People of any age at risk of acute kidney injury (AKI).

**Selection Criteria for Studies:** We systematically searched MEDLINE, Embase, Web of Science, and the Cochrane Controlled Trials registry from database inception to November 2023 for observational studies or RCTs that compared kidney outcomes among groups treated with cilastatin, either alone or as combination imipenem-cilastatin, versus an inactive or active control group not treated with cilastatin.

**Data Extraction:** Two reviewers independently evaluated studies for inclusionand risk of bias.

**Analytical Approach:** Treatment effects were estimated using random effects models and heterogeneity was quantified using the I^2^ statistic.

**Results:** We identified 10 studies (five RCTs, n=531 patients; 5 observational studies, n=6,321 participants) that met the inclusion criteria, including 6 studies with comparisons of imipenem-cilastatin to an inactive control and 4 studies with comparisons to alternate antibiotics. Based on pooled results from 5 studies, the risk of AKI was lower with imipenem-cilastatin (risk ratio [RR] 0.53 [95% CI, 0.38 to 0.74]; I^2^=42.2%), with consistent results observed from randomized trials (two trials, RR 0.30 [95% CI, 0.09 to 1.00]; I^2^=62.8%) and observational studies (three studies, RR 0.55 [95% CI, 0.38 to 0.81]; I^2^=62.8%). Based on results from six studies, kidney function was also better with imipenem-cilastatin than comparators (weighted mean difference [WMD] in serum creatinine -0.14 mg/dL [95% CI, -0.21 to -0.07]; I^2^=0%). The overall certainty of the evidence was low due to heterogeneity of the results, high risk of bias, and indirectness among the identified studies.

**Limitations:** Clinical and statistical heterogeneity could not be fully explained due to a limited number of studies.

**Conclusion:** Patients treated with imipenem-cilastatin developed AKI less frequently and had better short-term kidney function than control groups or those receiving comparator antibiotics. Larger clinical trials with less risk of detection bias due to lack of allocation concealment and blinding are needed to establish the efficacy of cilastatin for AKI prevention.

**Registration:** International Prospective Register of Systematic Reviews (PROSPERO) database (ID: CRD42023488809)

## BACKGROUND

Acute kidney injury (AKI) refers to a reduction in kidney function based on serum creatinine changes observed within 48 hours to 7 days or reduced urine output for 6 hours or more. ^1^ AKI is associated with increased mortality and the development of chronic conditions such as hypertension, cardiovascular disease, and chronic kidney disease (CKD). Complications from AKI result in higher healthcare costs and can also lead to reduced quality of life for affected individuals. ^2–6^

Several types of medications are have the potential to induce kidney tubular injury, thereby increasing the risks of acute and chronic kidney disease.. ^7,8^ Nephrotoxic AKI is most often caused by chemotherapeutic agents, antibiotics, calcineurin inhibitors, and radiocontrast dyes. ^7,9,10^ Hospitalized patients often receive these medications in the setting of acute illness, such as infection or at the time of procedures such as surgery or vascular procedures. ^11,12^ This places them at further risk of AKI, occurring in up to 25% of exposed patients, particularly among those with other health conditions. Effective strategies for preventing nephrotoxicity are needed to improve patient outcomes, reduce healthcare costs, and enhance quality of life.

Cilastatin was initially developed in the 1980s as an inhibitor of dehydropeptidase 1 (DPEP1) within the brush border of the renal proximal tubule to reduce the renal metabolism of imipenem, a broad-spectrum antibiotic prescribed for severe systemic infections. ^13–15^ By inhibiting DPEP1, cilastatin blocks the enzymatic hydrolysis of drugs before they are taken up into tubular epithelial cells where they can cause cell necrosis. ^16^ Cilastatin also blocks DPEP1 mediated leukocyte recruitment in the tubulointerstitial space, thereby reducing renal inflammation in response to injury. ^17^ Many animal studies have demonstrated nephroprotective effects of cilastatin, particularly following exposure to nephrotoxic drugs. ^18–20^ Specifically, cilastatin has been shown to reduce risk of kidney injury in rats following treatment with cyclosporin, imipenem, cisplatin, vancomycin, and radiocontrast dye. ^18,20–22^ Studies have also shown that cilastatin lowers the risk of kidney injury in rats undergoing kidney transplantation and in those receiving chemotherapeutic agents, without reducing the potency of the anticancer effect of these drugs. ^20,22^

The approval of imipenem-cilastatin for clinical use has enabled several studies in humans that suggest cilastatin may protect against drug-induced nephrotoxicity, including from fosfomycin, vancomycin, and cisplatin, ^23–25^ among patients undergoing solid organ transplantation, ^26–28^ bone marrow transplantation, ^29^ cancer therapy, ^30^ treatment of nosocomial pneumonia, ^31,32^ and childhood bacterial infections. ^33^ A previous meta-analysis of studies testing imipenem-cilastatin among kidney transplant recipients receiving cyclosporin reported better kidney function and lower incidence of AKI among patients who received imipenem-cilastatin when compared to a control group. ^34^ However, in the 16 years since that review, additional trials and comparative effectiveness studies have been published, suggesting that an updated systematic review and meta-analysis is needed to synthesize the current evidence base. ^35^ In this systematic review and meta-analysis, we examined the effects of cilastatin on AKI, kidney function, and subsequent clinical outcomes among people at risk of kidney injury.

## METHODS

We followed a pre-specified study protocol that was registered in the International Prospective Register of Systematic Reviews (PROSPERO) database (ID: CRD42023488809) ^36^ and adhered to the Preferred Reporting for Systematic Review and Meta-analysis (PRISMA) guidelines. ^37^

### Search Strategy

We conducted a comprehensive search using four electronic bibliographic databases including MEDLINE via OVID (from January 1946 to November 21, 2023), Embase via OVID (from January 1974 to November 2023), Web of Science (from January 1976 to November 22, 2023), and the Cochrane Controlled Trials Registry (from January 1996 to November 21, 2023). We developed the search strategy with the guidance of a health sciences librarian proficient in systematic search methodology. We used the following search terms as Medical Subject Heading (MeSH) and keyword combined with Boolean operators for the bibliographic database search; Mesh terms “Acute kidney injury,” OR “chronic kidney failure,”; Key heading word/Text word: “Acute kidney injury” OR “kidney injury” OR “renal injury” OR “renal insufficiency” OR “ kidney insufficiency” OR “chronic kidney injury” OR “end-stage kidney disease” OR “end-stage renal disease” OR “renal failure” or “kidney failure” OR “end-stage kidney failure” OR “end-stage renal failure” OR “kidney dysfunction” OR “nephroprotection” OR “nephrotoxicity” OR “all-cause mortality” OR “kidney function” OR “Creatinine” or “Cystatin C” or “glomerular filtration rate”, “urine output”, “allograft function” OR “proteinuria” or “albuminuria” or “kidney biomarkers” AND “Cilastatin” OR “cilastatin, imipenem drug combination.” We limited the search to studies in humans and included randomized controlled trials (RCTs) as well as comparative effectiveness observational study designs. There were no restrictions imposed on age, or language of publications. We excluded publications that were not primary research studies (e.g., editorials, narrative reviews, opinion pieces, letters, and research protocols, etc.). Additionally, citations and reference lists from included studies were also searched to identify other potentially relevant studies. The detailed literature search strategy for each electronic database is provided in the supplement (Table S1).

### Study Selection

Studies were eligible for inclusion if the population included human participants of any age at risk of AKI, AKD, or CKD arising from acute illness (e.g., infection, malignancy), medical or surgical procedures (e.g., transplant), or nephrotoxic exposures. Eligible studies were those including treatment with cilastatin either alone or in combination with imipenem and included a comparator group not treated with cilastatin; this could include an inactive control group with or without a placebo, or one or more active comparator groups not receiving cilastatin. Studies were included if they reported one or more outcomes of interest related to nephrotoxicity including a measure of kidney function (e.g., urine output, serum creatinine, cystatin C, measured or estimated glomerular filtration rate using any technique), kidney structure (e.g., albuminuria/proteinuria, abnormal urine sediment, kidney injury biomarkers including markers of tubular damage such as neutrophil gelatinase-associated lipocalin (NGAL), kidney injury molecule-1 (KIM-1), interleukin-18 (IL-18), kidney imaging, or kidney biopsy features), at risk of AKI based on serum creatinine changes or urine output criteria aligned with the Kidney Disease: Improving Global Outcomes (KDIGO), Acute Kidney Injury Network (AKIN), or risk, injury, failure, loss, end-stage kidney disease (RIFLE) criteria, ^38^ or as defined by the study authors. Additional outcomes of interest included downstream clinical outcomes of AKI, including all-cause mortality, development or progression of CKD, kidney failure, and cardiovascular events.

### Screening

We conducted a two-staged screening process to assess each article’s suitability for inclusion in our review. During the first stage of screening, each article’s title and abstract were independently reviewed by two authors (DA and FG). If there was uncertainty regarding inclusion based on the title and abstract alone by either reviewer, the article was retained for full-text review. Subsequently, a full-text review of all articles identified from the first stage was undertaken independently by the same two authors. In case of any disagreements arising among the reviewers at each screening stage, consensus was sought, and remaining disagreements were resolved by a third reviewer (MJ). To effectively organize the identified literature, we used Endnote 21 reference management software (Clarivate Analytics in Philadelphia, USA). ^39^

### Data extraction

A data extraction template was developed to systematically compile information from each eligible study. The data extraction process was distinct based on study design; a) RCTs, and b) comparative observational studies. Two authors (DA and FG) completed the data extraction from all studies. The specific data elements acquired included the primary author names, year of publication, geographical origin, study design, sample size, nature of the study population, participant age, sex distribution, and the documented study outcomes and their definitions. We sought to preferentially use definitions of AKI that aligned with the KDIGO, AKIN, RIFLE criteria ^38^ where possible, but used the definition provided by the study authors if the former were not reported. For studies where measures of kidney function were taken at multiple time points, we used the results from the last time point reported up to 90 days to define short-term changes and measurements after 90 days to identify long-term kidney function from each study.

### Risk of bias assessment

We assessed the risk of bias of each study using the Cochrane risk of bias tool for randomized trials (RoB tool version 2) ^40^ and the JBI critical appraisal tool for observational studies. ^41^ Each study underwent evaluation and was categorized into one of three levels of risk of bias: low risk, unclear risk, or high risk of bias.

### Statistical analyses

We quantified the agreement on article eligibility between reviewers in the first and second stages of article selection using the kappa (κ) statistic. The decision to perform meta-analysis was contingent upon the availability of at least two studies that met our predefined study inclusion criteria for each outcome and that were considered clinically similar enough to justify pooling results.

Given expected clinical and statistical heterogeneity between studies, we estimated pooled dichotomous outcomes using random effects models according to the DerSimonian and Laird method, ^42^ with treatment effects estimated as risk ratios (RRs) with 95% confidence intervals (Cis). Continuous outcomes were also pooled using random effects models incorporating restricted maximum likelihood (REML) weighting to estimate weighted mean differences with 95% CIs. ^43^ Between-study heterogeneity was assessed using the I^2^ statistic. We conducted pre-specified subgroup analyses and meta-regression for each outcome according to study design (RCT or observational study). Publication bias was investigated using funnel plots and Egger’s test. ^44,45^ The statistical significance threshold for all tests was set at p<0.05. Analyses were conducted using Stata Statistical Software, StataCorp version 17 (Stata Corporation, College Station, TX, USA), using the ‘metan’ package. ^46^

### Assessment of certainty of the evidence

The certainty of evidence was evaluated by two authors (DA and MJ) using the Grading of Recommendations, Assessment, Development and Evaluations (GRADE) approach to determine whether the overall certainty of the evidence for the nephroprotective effects of cilastatin in humans as: very low, low, moderate, or high. ^47^

## RESULTS

### Selection of studies

The electronic database search yielded 1,015 citations. Among these, 190 citations were identified as duplicates and removed. In the first stage of screening, 732 articles were excluded based on title and abstract, resulting in 93 articles selected for full-text review. Of these, 10 studies were identified that met the inclusion criteria. There was a high level of agreement between reviewers in the selection of articles for inclusion (kappa statistic, κ =0.61). The study selection process is represented in further detail in the PRISMA flowchart (Figure 1). ^48^

**Figure 1.**
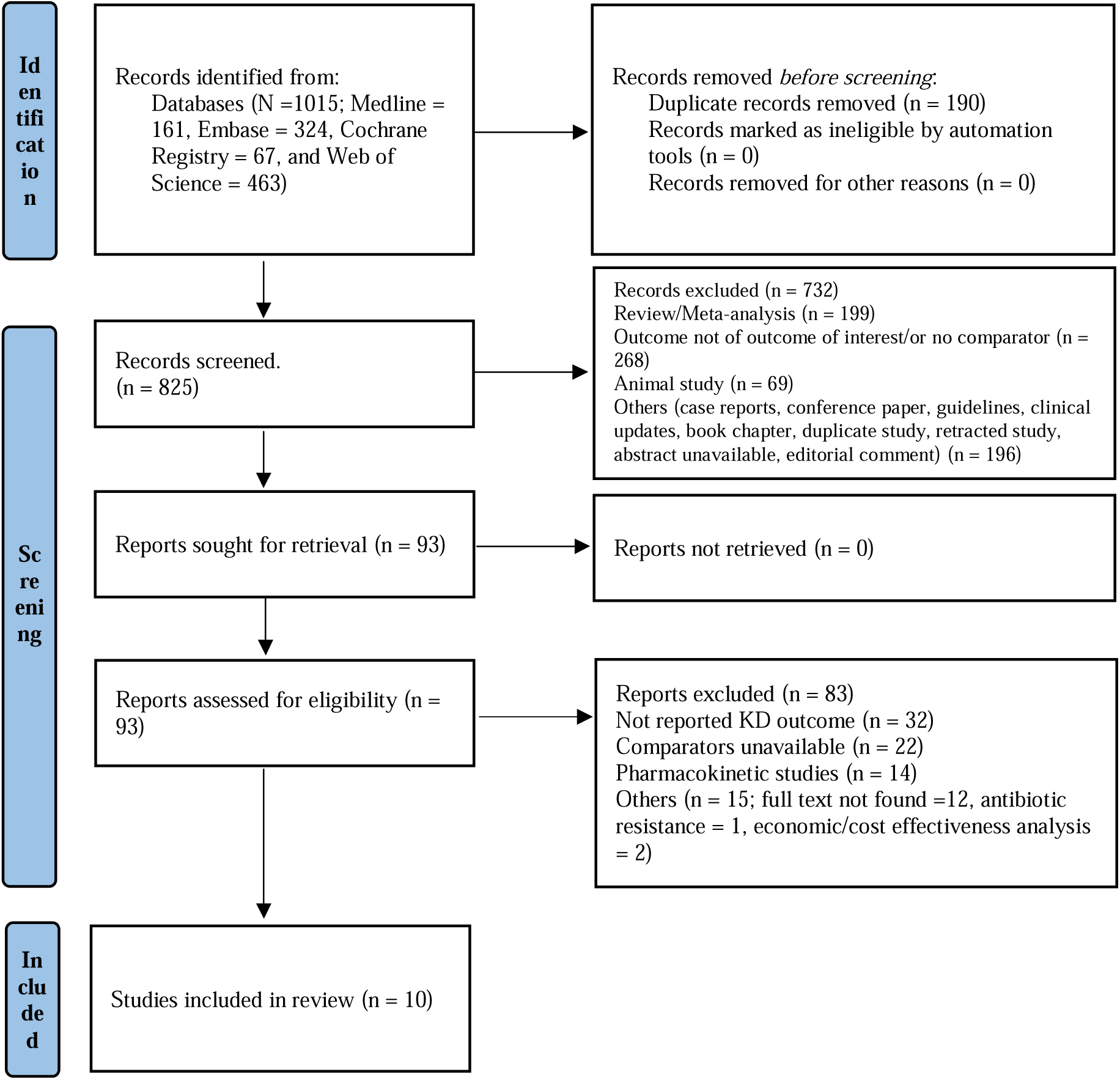
PRISMA flow diagram

### Study characteristics

The characteristics of the included studies are reported in Table 1. Of the 10 included studies, five were RCTs ^27,28,31,32,49^ (n=531 patients), and five were observational comparative effectiveness studies ^26,29,30,33,50^ (n=6,321 participants). Publication dates ranged from 1994 to 2021, and the number of participants per study varied from 20 to 5,566. Study populations included kidney transplant recipients (2 studies), ^27,28^ heart and lung transplant recipients (1 study), ^26^ bone marrow transplant recipients (1 study), ^29^ patients treated for nosocomial infections (2 studies), ^31,32^ heart transplant recipients (1 study), ^49^ infants with severe bacterial infection (1 study), ^33^ and patients receiving chemotherapy for peritoneal carcinomatosis (1 study). ^30^ All studies tested imipenem/cilastatin as the intervention. The comparison groups varied across studies, with an inactive control used in 6 studies (with one describing use of a placebo control) ^26–30,49^ and an active comparator used in the other 4 studies, including meropenem in two studies, ^33,50^ piperacillin/tazobactam in one study, ^31^ and cefepime in one study. ^32^

**Table 1.**
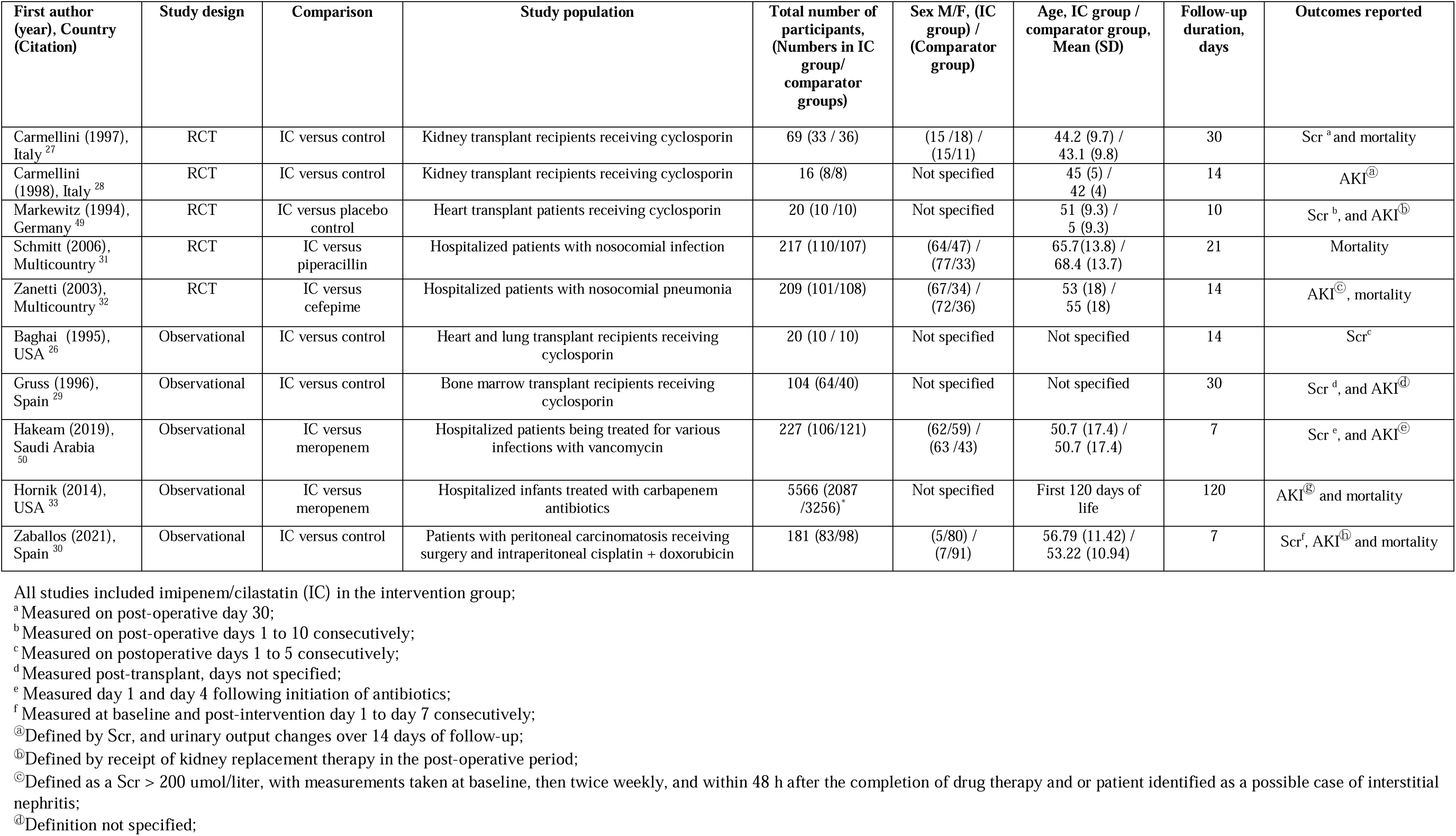
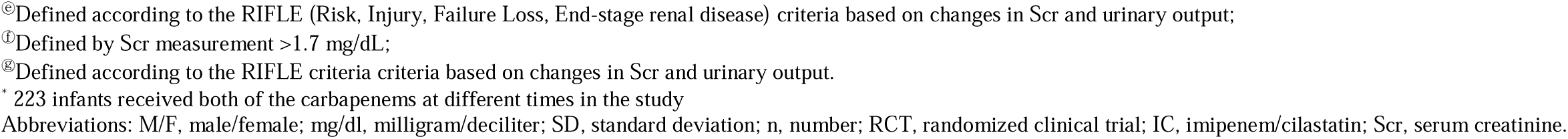
Characteristics of the included studies.

| First author (year), Country (Citation) | Study design | Comparison | Study population | Total number of participants, (Numbers in IC group/ comparator groups) | Sex M/F, (IC group) / (Comparator group) | Age, IC group / comparator group, Mean (SD) | Follow-up duration, days | Outcomes reported |
| --- | --- | --- | --- | --- | --- | --- | --- | --- |
| Carmellini (1997), Italy<sup>27</sup> | RCT | IC versus control | Kidney transplant recipients receiving cyclosporin | 69 (33 / 36) | (15 / 18) / (15/11) | 44.2 (9.7) / 43.1 (9.8) | 30 | Scr<sup>a</sup> and mortality |
| Carmellini (1998), Italy<sup>28</sup> | RCT | IC versus control | Kidney transplant recipients receiving cyclosporin | 16 (8/8) | Not specified | 45 (5) / 42 (4) | 14 | AKI<sup>®</sup> |
| Markewitz (1994), Germany<sup>49</sup> | RCT | IC versus placebo control | Heart transplant patients receiving cyclosporin | 20 (10 / 10) | Not specified | 51 (9.3) / 5 (9.3) | 10 | Scr<sup>b</sup>, and AKI<sup>®</sup> |
| Schmitt (2006), Multicountry<sup>31</sup> | RCT | IC versus piperacillin | Hospitalized patients with nosocomial infection | 217 (110/107) | (64/47) / (77/33) | 65.7(13.8) / 68.4 (13.7) | 21 | Mortality |
| Zanetti (2003), Multicountry<sup>32</sup> | RCT | IC versus cefepime | Hospitalized patients with nosocomial pneumonia | 209 (101/108) | (67/34) / (72/36) | 53 (18) / 55 (18) | 14 | AKI<sup>®</sup>, mortality |
| Baghai (1995), USA<sup>26</sup> | Observational | IC versus control | Heart and lung transplant recipients receiving cyclosporin | 20 (10 / 10) | Not specified | Not specified | 14 | Scr<sup>c</sup> |
| Gruss (1996), Spain<sup>29</sup> | Observational | IC versus control | Bone marrow transplant recipients receiving cyclosporin | 104 (64/40) | Not specified | Not specified | 30 | Scr<sup>d</sup>, and AKI<sup>®</sup> |
| Hakeam (2019), Saudi Arabia<sup>50</sup> | Observational | IC versus meropenem | Hospitalized patients being treated for various infections with vancomycin | 227 (106/121) | (62/59) / (63 /43) | 50.7 (17.4) / 50.7 (17.4) | 7 | Scr<sup>e</sup>, and AKI<sup>®</sup> |
| Hornik (2014), USA<sup>33</sup> | Observational | IC versus meropenem | Hospitalized infants treated with carbapenem antibiotics | 5566 (2087 /3256)<sup>g</sup> | Not specified | First 120 days of life | 120 | AKI<sup>®</sup> and mortality |
| Zaballos (2021), Spain<sup>30</sup> | Observational | IC versus control | Patients with peritoneal carcinomatosis receiving surgery and intraperitoneal cisplatin + doxorubicin | 181 (83/98) | (5/80) / (7/91) | 56.79 (11.42) / 53.22 (10.94) | 7 | Scr<sup>f</sup>, AKI<sup>®</sup> and mortality |

All studies included imipenem/cilastatin (IC) in the intervention group;
<sup>a</sup> Measured on post-operative day 30;
<sup>b</sup> Measured on post-operative days 1 to 10 consecutively;
<sup>c</sup> Measured on postoperative days 1 to 5 consecutively;
<sup>d</sup> Measured post-transplant, days not specified;
<sup>e</sup> Measured day 1 and day 4 following initiation of antibiotics;
<sup>f</sup> Measured at baseline and post-intervention day 1 to day 7 consecutively;
<sup>g</sup> Defined by Scr, and urinary output changes over 14 days of follow-up;
<sup>®</sup> Defined by receipt of kidney replacement therapy in the post-operative period;

Outcomes of interest included AKI reported in five studies using varying definitions, ^28–30,33,49^ short-term changes in serum creatinine reported in six studies with the last time point of measurement ranging from 5 to 30 days of follow-up,^26,27,29,30,49,50^ and all-cause mortality reported in five studies.^27,30–33,50^ We identified no studies examining the outcomes of development or progression of CKD, long-term kidney function, kidney failure, or cardiovascular events.

### Effect of imipenem-cilastatin on acute kidney injury

Based on results from five studies ^28–30,33,49^ including a total of 6,074 participants, those treated with imipenem-cilastatin had a lower risk of AKI compared to comparators (pooled RR 0.53 [95% CI, 0.38 to 0.74]), with moderate heterogeneity observed between studies (I^2^=42.2%).

Treatment effects of imipenem-cilastatin on AKI were consistent by study design (meta-regression p-value=0.35); among 2 RCTs the pooled RR was 0.30 (95% CI, 0.09 to 1.00), I^2^=68.2%, while among 3 observational studies the pooled RR was 0.55 (95% CI, 0.38 to 0.81), I^2^=68.2% (Figure 2).

**Figure 2.**
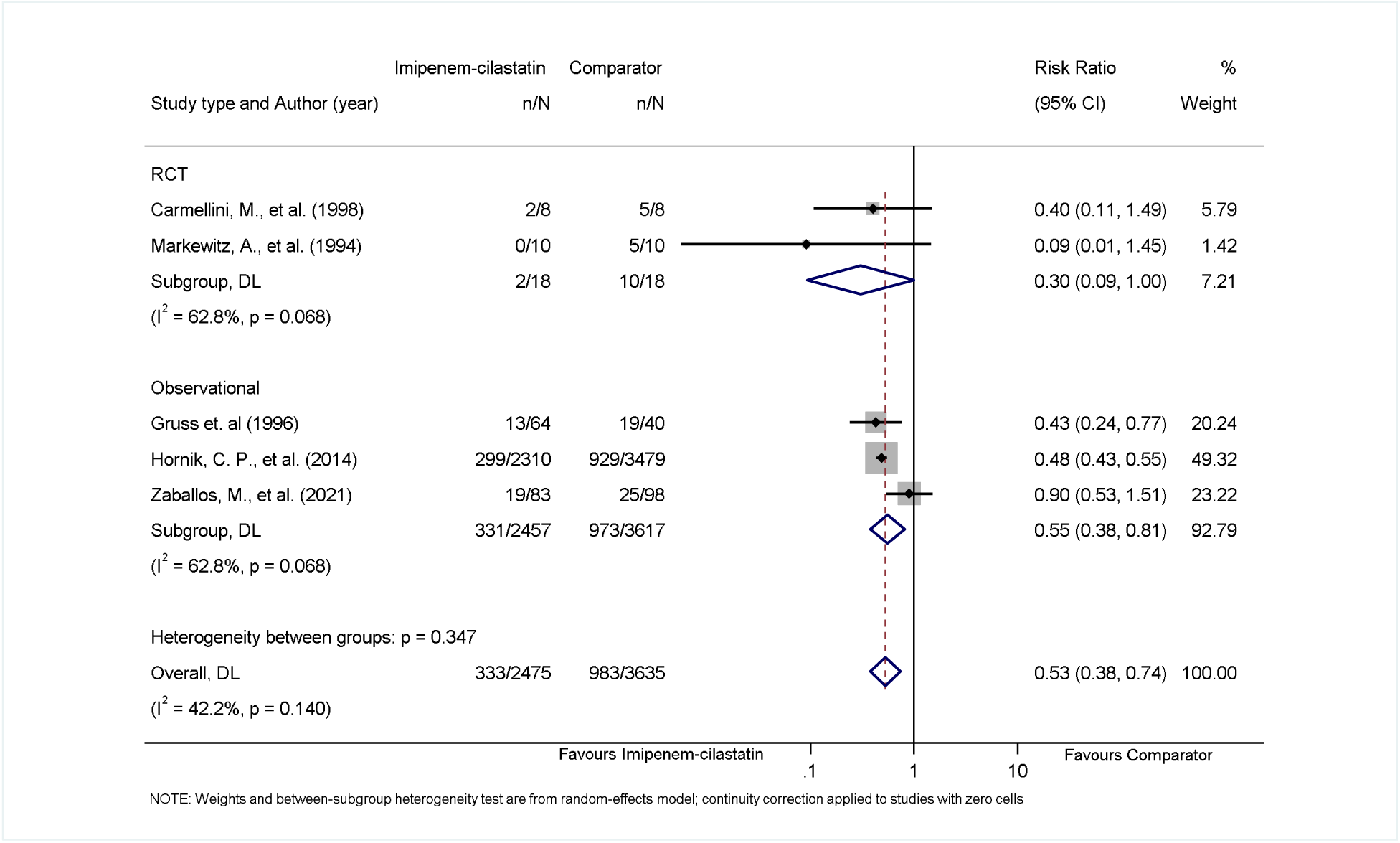
Forest plot demonstrating pooled effect of imipenem-cilastatin on acute kidney injury from all studies and stratified by randomized controlled trial or observational study design. Note: The study by Hornik et al included 223 infants who received imipenem-cilastatin and the comparator carbapenem antibiotics at different times. Abbreviations: CI, confidence interval; DL, DerSimonian and Laird; n, number of acute kidney injury events; N, total number of study participants; RCT, randomized controlled trial.

### Effect of imipenem-cilastatin on kidney function

Results from 6 studies showed that patients treated with imipenem-cilastatin had better short-term kidney function compared to comparators;^26,27,29,30,49,50^ weighted mean difference in serum creatinine was -0.14 mg/dL (95% CI, -0.21 to -0.07) with no evidence of statistical heterogeneity observed between studies (I^2^=0%). Results remained consistent between RCTs and observational studies (meta-regression p=0.46) (Figure 3).

**Figure 3.**
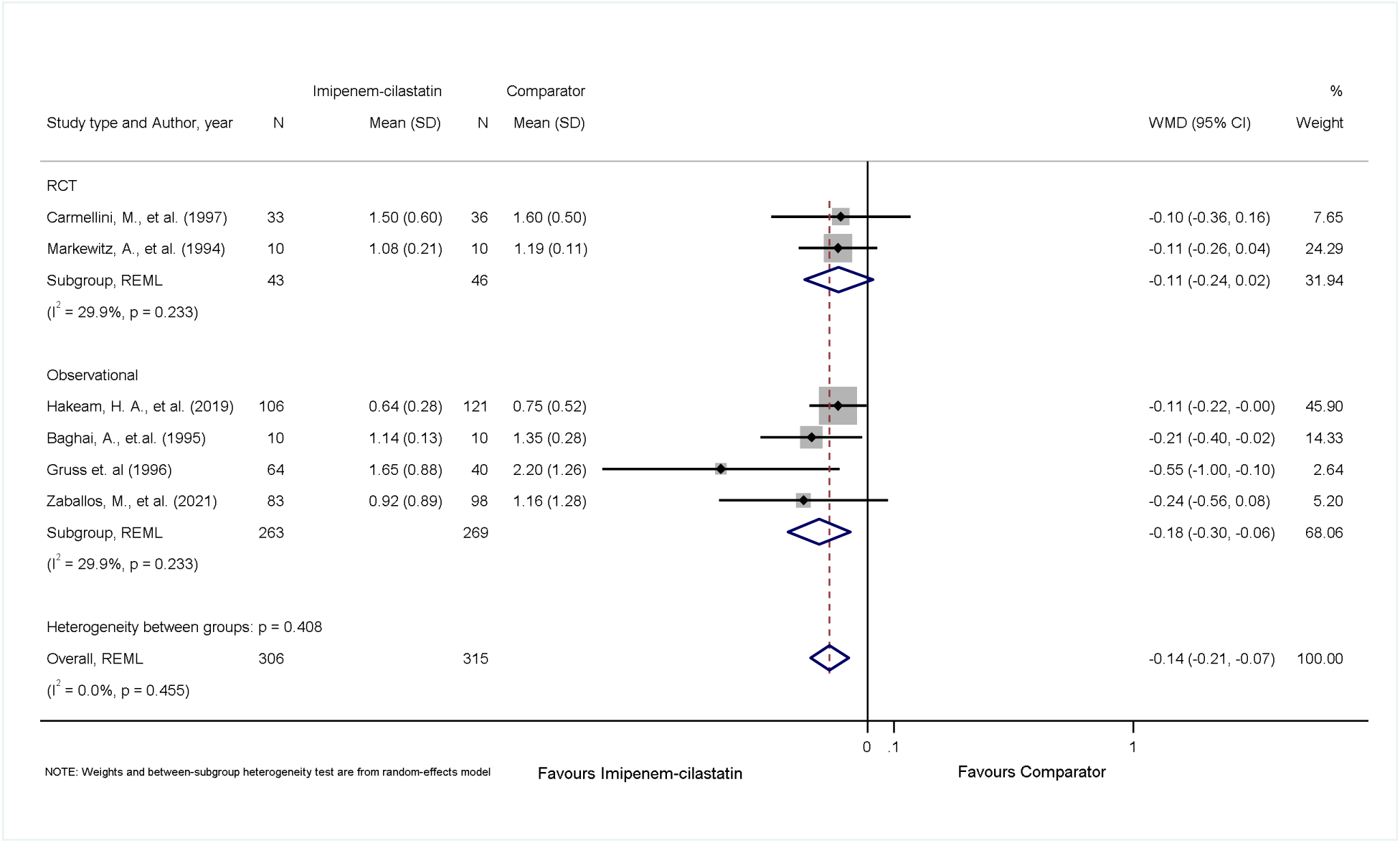
Forest plot demonstrating pooled effect of imipenem-cilastatin on serum creatinine from all studies and stratified by randomized controlled trial or observational study design. Abbreviations: CI, confidence interval; N, total number of study participants in imipenem-cilastatin or comparator group for individual study; RCT, randomized controlled trial; REML, restricted maximum likelihood; SD, standard deviation; WMD, weighted mean difference.

### Effect of imipenem-cilastatin on all-cause mortality

Six studies reported on all-cause mortality.^27,30–33,50^ Patients treated with imipenem-cilastatin experienced no statistically significant difference in all-cause mortality compared with comparators (pooled RR 0.82 [95% CI, 0.44 to 1.54] with a high degree of heterogeneity across studies (I^2^=74.2%) (Figure 4). Pooled estimates were consistent between RCTs and observational studies (meta-regression p=0.916).

**Figure 4.**
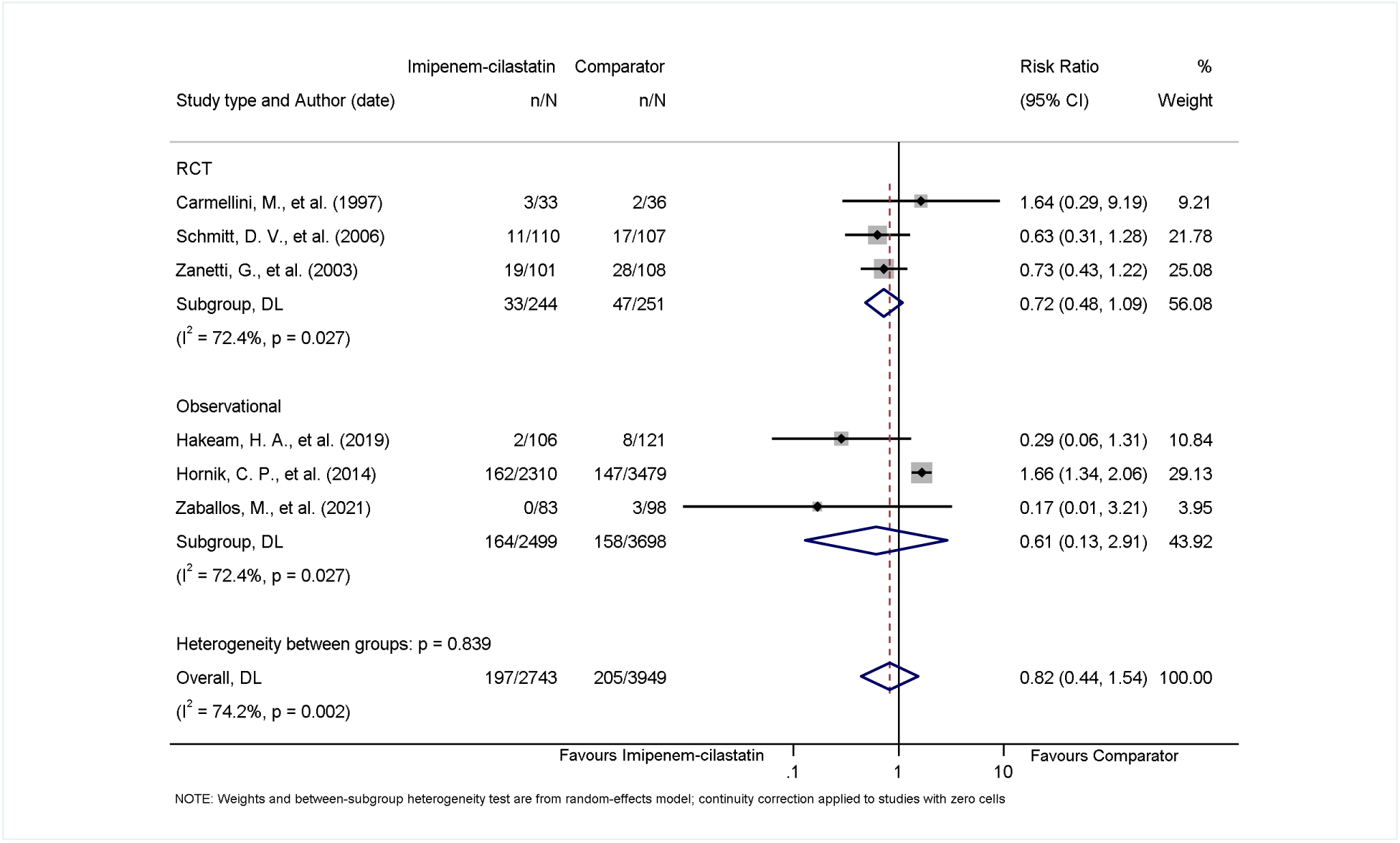
Forest plot demonstrating pooled effect of imipenem-cilastatin on all-cause mortality from all studies and stratified by randomized controlled trial or observational study design. Note: The study by Hornik et al included 223 infants who received imipenem-cilastatin and the comparator carbapenem antibiotics at different times. Abbreviations: CI, confidence interval; DL, DerSimonian and Laird; n, number of acute kidney injury events; N, total number of study participants; RCT, randomized controlled trial.

### Risk of bias

The risk of bias of RCTs according to the RoB 2.0 tool is shown in Table 2. Four of the five trials were at unclear or high risk of bias due to lack of or unclear allocation concealment. All trials were at high or unclear risk of detection bias due to lack of or unclear blinding, and three were at unclear risk of attrition bias due to lack of reporting of losses to follow-up. The risk of bias of observational studies according to the JBI critical appraisal tool for observational studies is shown in Table 3. Three of the five observational studies were at unclear or high risk of bias due to unclear or inadequate strategies to address confounding.

**Table 2.**
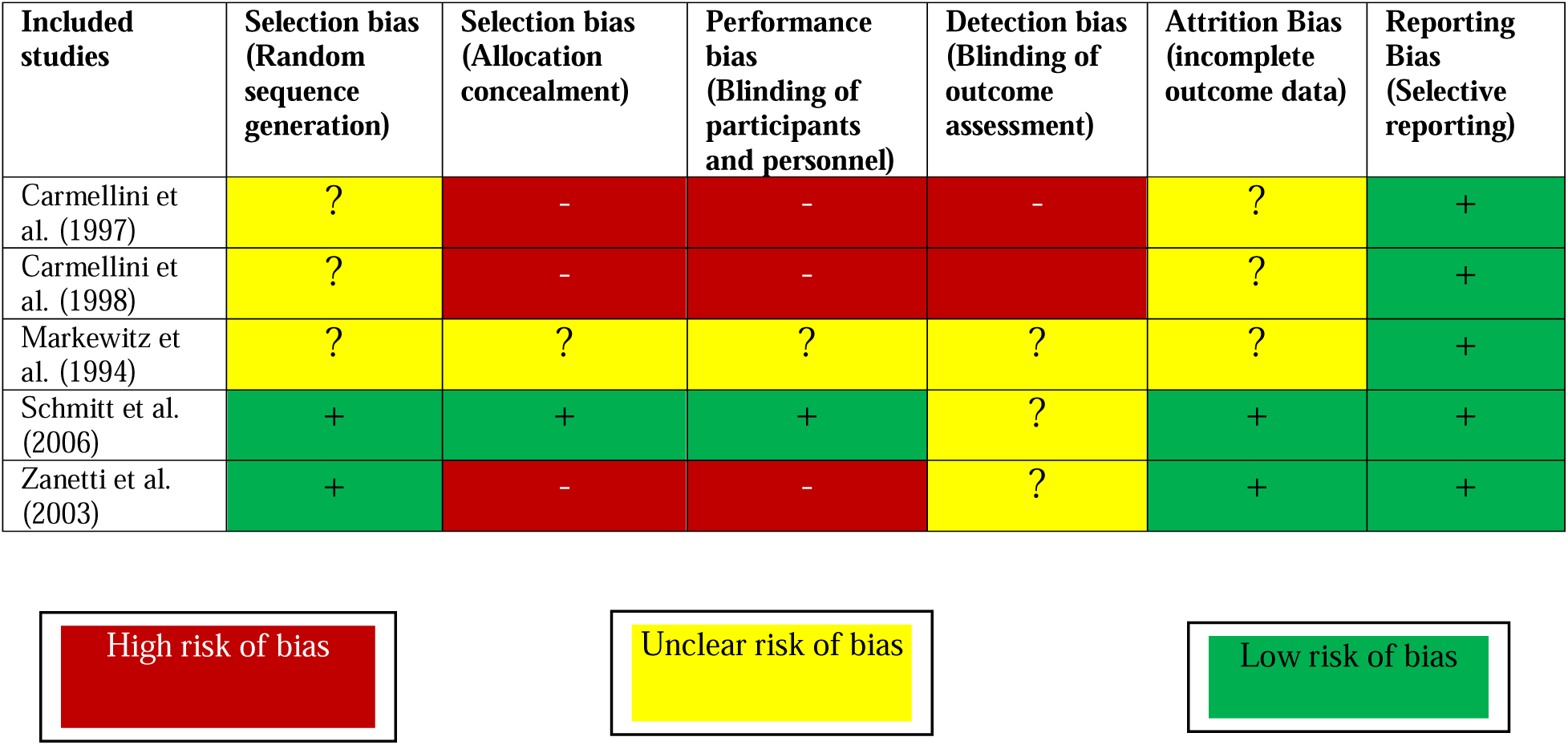
Risk of bias of randomized trials of imipenem-cilastatin adapted from the Cochrane Risk of Bias tool (RoB tool version 2)

**Table 3.** Risk of bias assessment of observational studies of imipenem-cilastatin adapted from the Joanne Briggs Institute critical appraisal checklist.

| Included studies | Criteria for inclusion in the sample clearly defined | Subjects and the setting described in detail | Exposure measured in a valid and reliable way | Objective, standard criteria used for measurement of the condition | Confounding factors identified | Strategies to deal with confounding factors stated | Outcomes measured in a valid and reliable way | Appropriate statistical analysis used |
| --- | --- | --- | --- | --- | --- | --- | --- | --- |
| Baghai et al. (1995) | + | - | + | + | - | - | + | ? |
| Gruss et al. (1996) | + | ? | + | ? | - | - | ? | + |
| Hakeam et al. (2019) | + | + | + | + | + | + | + | + |
| Hornik et al. (2014) | + | + | + | + | + | + | + | + |
| Zaballos et al. (2021) | + | + | + | + | ? | ? | + | + |

High risk of bias
Unclear risk of bias
Low risk of bias

### Publication bias

Funnel plots for AKI, serum creatinine, and mortality showed asymmetry in-keeping with small study effects suggestive of publication bias (Supplementary Figure S1). However, there was no statistical evidence of small study effects based on Egger’s test for the outcomes of AKI (p=0.342), serum creatinine (p=0.079), or all-cause mortality (p=0.093), although the number of studies limited the power of these tests.

### Certainty of the Evidence

The overall certainty of the evidence was graded as low due to moderate and high statistical heterogeneity for the outcomes of AKI and mortality, high risk of bias for most of the individual studies, and indirectness (use of surrogate outcomes).

## DISCUSSION

In this systematic review and meta-analysis, we evaluated the effects of cilastatin compared to inactive or active comparators, on kidney outcomes. We identified 5 RCTs and 5 observational studies that evaluated effects on the outcomes of AKI, kidney function (based on serum creatinine), and all-cause mortality among human participants at risk of AKI in a variety of clinical settings. All studies included in our review used cilastatin in combination with imipenem. In our meta-analysis, we found that imipenem-cilastatin reduced the risk of developing AKI by 49%, although there was heterogeneity in the effect size between studies. We also found that serum creatinine measured after short-term follow-up (range 5-30 days) was better among participants who received imipenem-cilastatin than comparators. There was no statistically significant difference in all-cause mortality among participants who received imipenem-cilastatin compared with comparators. For all outcomes, results were similar among RCTs and observational studies. It is important to note that many of the studies were at high risk of bias, and funnels plots suggested that publication bias may exist.

The nephroprotective effects of cilastatin have been demonstrated in several pre-clinical studies ^18–22^ and mechanistic effects have been further examined in a number of human studies. ^23–29^ The mechanism of action for cilastatin involves counteracting metabolism of nephrotoxic substances in the proximal tubule of the kidney, thereby blocking the uptake of these agents and preventing tubular necrosis through several pathways, including via reactive oxygen species, inflammation, and apoptosis. In addition to reducing nephrotoxin accumulation in renal tubular cells, cilastatin is believed to attenuate leukotriene mediated interstitial inflammation. ^24,51,52^ The reported nephroprotective effects of cilastatin from pre-clinical studies have generated interest in its use as an agent for prevention of AKI caused by nephrotoxic medication exposures, and the availability of imipenem-cilastatin formulation has enabled comparative studies that have evaluated kidney outcomes in several clinical settings. ^35^

Our study findings align with those from a previous meta-analysis conducted by Tejedor et al. in 2007, ^34^ which included five studies among patients with organ transplantation receiving imipenem-cilastatin compared to cyclosporine. They also reported lower serum creatinine concentrations for patients treated with imipenem-cilastatin and a 76% reduction in the odds of developing acute renal failure. Our updated review identified 5 additional studies published since their review and provides an updated and consolidated evidence base including additional studies with comparisons to alternative antibiotics demonstrating nephroprotective effects of imipenem-cilastatin. In this review, the study participants were from diverse clinical settings, where they were exposed to pharmacological agents or procedures that confer risk of AKI. These interventions involved antibiotic therapy for bacterial infections, cancer chemotherapy, as well as the administration of calcineurin inhibitor in patients undergoing solid organ transplantation, suggesting cilastatin might be a AKI preventive approach that could be applied in several clinical settings.

This review has several limitations that are important to acknowledge. First, the studies included had a large degree of clinical heterogeneity, not only in the clinical populations and settings, but also in the way outcomes were measured, including the definition used for AKI and the timing and methodology of serum creatinine measurement. Studies did not consistently define AKI using the KDIGO definition, requiring the use of a variety of definitions as reported by the authors that differed in their incorporation of serum creatinine thresholds, incorporation of urine output, and identification of treatment with dialysis across the studies. Furthermore, measurements of kidney function were made at different time points after treatment in these studies, and all were within short-term periods of follow-up. We were able to address this in our meta-analysis by selecting the last available serum creatinine reported by each study; however, this makes interpretation of the pooled difference challenging since kidney function may vary (with both worsening and improvement) with time following AKI. Second, the number of studies identified was small, and the existing RCTs that were identified had small sample sizes and were at high risk of bias. The small number of studies limited our ability to explore reasons for statistical heterogeneity and detect publication bias. Third, the comparator groups varied across studies, with a number including an active comparator including an alternative broad-spectrum antibiotic. It is thus possible that these comparisons are confounded by differences in the risk of AKI with imipenem versus meropenem or beta-lactam antibiotics, rather than being attributable to an independent effect of cilastatin itself. We did not identify trials of cilastatin alone, which may have a more favourable safety and efficacy profile that imipenem-cilastatin if used for AKI prevention alone. New trials testing formulations of cilastatin for AKI prevention are needed to test this hypothesis. Finally, the studies identified largely relied on surrogate endpoints such as AKI and serum creatinine differences, rather than patient-centred clinical outcomes such as major adverse kidney outcomes.

In conclusion, this systematic review and meta-analysis suggests that cilastatin may reduce the risk of AKI; however, the existing evidence base is derived from studies of imipenem-cilastain with a high risk of bias and efficacy is uncertain due to the statistical heterogeneity of findings, indirectness of the evidence base, and potential detection and publication biases. Further large-scale randomized placebo-controlled trials of cilastatin with appropriate allocation concealment and blinding and focused on clinically important outcomes are needed to determine the efficacy and safety of cilastatin used for AKI prevention among patients receiving contemporary nephrotoxic exposures.

## Data Availability

All data produced in the present study are available upon reasonable request to the authors.

## ACKNOWLEDGEMENT

The authors thank Diane L. Lorenzetti, from the Health Sciences Library, University of Calgary for her assistance in developing the electronic bibliographic database search strategy.

## DATA SHARING

All data produced in the present study are available upon reasonable request to the authors.

## REFERENCES

1. Makris K, Spanou L. Acute Kidney Injury: Definition, Pathophysiology and Clinical Phenotypes. Clin Biochem Rev. May 2016;37(2):85–98.

2. Zhang S, Ren H-F, Du R-X, Sun W-L, Fu M-L, Zhang X-C. Global, regional, and national burden of kidney dysfunction from 1990 to 2019: a systematic analysis from the global burden of disease study 2019. BMC Public Health. 2023/06/23 2023;23(1):1218. doi:10.1186/s12889-023-16130-8

3. Sarnak MJ, Levey AS, Schoolwerth AC, et al. Kidney Disease as a Risk Factor for Development of Cardiovascular Disease. Circulation. 2003;108(17):2154–2169. doi:doi:10.1161/01.CIR.0000095676.90936.80

4. Saran R, Pearson A, Tilea A, et al. Burden and Cost of Caring for US Veterans With CKD: Initial Findings From the VA Renal Information System (VA-REINS). American Journal of Kidney Diseases. 2021/03/01/ 2021;77(3):397–405. doi:10.1053/j.ajkd.2020.07.013

5. Matsushita K, van der Velde M, Astor BC, et al. Association of estimated glomerular filtration rate and albuminuria with all-cause and cardiovascular mortality in general population cohorts: a collaborative meta-analysis. Lancet. Jun 12 2010;375(9731):2073–81. doi:10.1016/s0140-6736(10)60674-5

6. Matsushita K, Coresh J, Sang Y, et al. Estimated glomerular filtration rate and albuminuria for prediction of cardiovascular outcomes: a collaborative meta-analysis of individual participant data. Lancet Diabetes Endocrinol. Jul 2015;3(7):514–25. doi:10.1016/s2213-8587(15)00040-6

7. Awdishu L, Mehta RL. The 6R’s of drug induced nephrotoxicity. BMC Nephrology. 2017/04/03 2017;18(1):124. doi:10.1186/s12882-017-0536-3

8. Sharma V, Singh TG. Drug induced nephrotoxicity-A mechanistic approach. Molecular Biology Reports. 2023/08/01 2023;50(8):6975–6986. doi:10.1007/s11033-023-08573-4

9. Kane-Gill SL, Goldstein SL. Drug-Induced Acute Kidney Injury: A Focus on Risk Assessment for Prevention. Critical Care Clinics. 2015/10/01/ 2015;31(4):675–684. doi:10.1016/j.ccc.2015.06.005

10. Mendoza SA. Nephrotoxic drugs. Pediatr Nephrol. Oct 1988;2(4):466–76. doi:10.1007/bf00853443

11. Patel JB, Sapra A. Nephrotoxic medications. 2020;

12. Perazella MA, Rosner MH. Drug-Induced Acute Kidney Injury. Clin J Am Soc Nephrol. Aug 2022;17(8):1220–1233. doi:10.2215/cjn.11290821

13. Calandra GB, Ricci FM, Wang C, Brown KR. The Efficacy Results and Safety Profile of Imipenem/Cilastatin from the Clinical Research Trials. The Journal of Clinical Pharmacology. 1988;28(2):120–127. doi:10.1002/j.1552-4604.1988.tb05735.x

14. Drusano GL, Standiford HC. Pharmacokinetic profile of lmipenem/cilastatin in normal volunteers. The American Journal of Medicine. 1985/06/07/ 1985;78(6, Supplement 1):47–53. doi:10.1016/0002-9343(85)90101-9

15. Kahan FM, Kropp H, Sundelof JG, Birnbaum J. Thienamycin: development of imipenem-cilastatin. Journal of Antimicrobial Chemotherapy. 1983;12(suppl_D):1–35. doi:10.1093/jac/12.suppl_D.1

16. Buckley MM, Brogden RN, Barradell LB, Goa KL. Imipenem/Cilastatin. Drugs. 1992/09/01 1992;44(3):408–444. doi:10.2165/00003495-199244030-00008

17. Lau A, Rahn JJ, Chappellaz M, et al. Dipeptidase-1 governs renal inflammation during ischemia reperfusion injury. Science Advances. 2022;8(5):eabm0142. doi:doi:10.1126/sciadv.abm0142

18. Humanes B, Camaño S, Lara JM, et al. Cisplatin-induced renal inflammation is ameliorated by cilastatin nephroprotection. Article. Nephrol Dial Transplant. Oct 2017;32(10):1645–1655. doi:10.1093/ndt/gfx005

19. Humanes B, Jado JC, Camaño S, et al. Protective Effects of Cilastatin against Vancomycin-Induced Nephrotoxicity. Article. Biomed Res Int. 2015;2015:12. 704382. doi:10.1155/2015/704382

20. Humanes B, Lazaro A, Camano S, et al. Cilastatin protects against cisplatin-induced nephrotoxicity without compromising its anticancer efficiency in rats. Article. Kidney International. Sep 2012;82(6):652–663. doi:10.1038/ki.2012.199

21. Hammer C, Thies JC, Mraz W, Mihatsch M. Reduction of cyclosporine (csa) nephrotoxicity by imipenem cilastatin after kidney-transplantation in rats. Article. Transplant Proc. Feb 1989;21(1):931–931.

22. Sido B, Hammer C, Mraz W, Krombach F. Nephroprotective effect of imipenem cilastatin in reducing cyclosporine toxicity. Article. Transplant Proc. Feb 1987;19(1):1755–1758.

23. Im Ds, Shin Hj, Yang KJ, et al. Cilastatin attenuates vancomycin-induced nephrotoxicity via P-glycoprotein. Toxicology Letters. 2017/08/05/ 2017;277:9–17. doi:10.1016/j.toxlet.2017.05.023

24. Luo K, Lim SW, Jin J, et al. Cilastatin protects against tacrolimus-induced nephrotoxicity via anti-oxidative and anti-apoptotic properties. BMC Nephrol. Jun 14 2019;20(1):221. doi:10.1186/s12882-019-1399-6

25. Ortega-Trejo JA, Pérez-Villalva R, Arreola-Guerra JM, Ramírez V, Sifuentes-Osornio J, Bobadilla NA. Effect of Fosfomycin on Cyclosporine Nephrotoxicity. Antibiotics (Basel). Oct 21 2020;9(10)doi:10.3390/antibiotics9100720

26. Baghaie A BM, Abobo C, et al.,. The effect of imipenem/ cilastatin on acute cyclosporine nephrotoxicity in heart/lung transplant patients. 1995:23:A241.

27. Carmellini M, Frosini F, Filipponi F, Boggi U, Mosca F. Effect of cilastatin on cyclosporine-induced acute nephrotoxicity in kidney transplant recipients. Article. Transplantation. Jul 1997;64(1):164–166. doi:10.1097/00007890-199707150-00029

28. Carmellini M, Matteucci E, Boggi U, Cecconi S, Giampietro O, Mosca F. Imipenem/cilastatin reduces cyclosporin-induced tubular damage in kidney transplant recipients. Article; Proceedings Paper. Transplant Proc. Aug 1998;30(5):2034–2035. doi:10.1016/s0041-1345(98)00523-5

29. Gruss E, Tomas JF, Bernis C, Rodriguez F, Traver JA, FernandezRanada JM. Nephroprotective effect of cilastatin in allogeneic bone marrow transplantation. Results from a retrospective analysis. Article. Bone Marrow Transplant. Oct 1996;18(4):761–765.

30. Zaballos M, Power M, Canal-Alonso MI, et al. Effect of Cilastatin on Cisplatin-Induced Nephrotoxicity in Patients Undergoing Hyperthermic Intraperitoneal Chemotherapy. Article. Int J Mol Sci. Feb 2021;22(3):17. 1239. doi:10.3390/ijms22031239

31. Schmitt DV, Leitner E, Welte T, Lode H. Piperacillin/tazobactam vs imipenem/cilastatin in the treatment of nosocomial pneumonia - a double blind prospective multicentre study. Article. Infection. Jun 2006;34(3):127–134. doi:10.1007/s15010-006-5020-0

32. Zanetti G, Bally F, Greub G, et al. Cefepime versus imipenem-cilastatin for treatment of nosocomial pneumonia in intensive care unit patients: A multicenter, evaluator-blind, prospective, randomized study. Article. Antimicrob Agents Chemother. Nov 2003;47(11):3442–3447. doi:10.1128/aac.47.11.3442-3447.2003

33. Hornik CP, Herring AH, Benjamin DK, Jr., et al. Adverse events associated with meropenem versus imipenem/cilastatin therapy in a large retrospective cohort of hospitalized infants. Comparative Study Research Support, N.I.H., Extramural Research Support, Non-U.S. Gov’t. Pediatric Infectious Disease Journal. 2014;32(7):748–53. doi:10.1097/INF.0b013e31828be70b

34. Tejedor A, Torres AM, Castilla M, Lazaro JA, Lucas Cd, Caramelo C. Cilastatin protection against cyclosporin A-induced nephro-toxicity: clinical evidence. Current Medical Research and Opinion. 2007/03/01 2007;23(3):505–513. doi:10.1185/030079906X167633

35. Shayan M, Elyasi S. Cilastatin as a protective agent against drug-induced nephrotoxicity: a literature review. Expert Opin Drug Saf. Aug 2020;19(8):999–1010. doi:10.1080/14740338.2020.1796967

36. Acharya D, Ghanim F, Harrison T, Ronksely P, Elliott MJ, Collister D, Scory TD, Shommu N, Pannu N, James MT. Nephroprotective Effects of Cilastatin in People at Risk of Acute Kidney Injury: Systematic Review and Meta-analysis. 2023 (available from: https://www.crd.york.ac.uk/prospero/display_record.php?ID=CRD42023488809)

37. Page MJ, McKenzie JE, Bossuyt PM, et al. The PRISMA 2020 statement: an updated guideline for reporting systematic reviews. BMJ. 2021;372:n71. doi:10.1136/bmj.n71

38. Yaqub S, Hashmi S, Kazmi MK, Aziz Ali A, Dawood T, Sharif H. A Comparison of AKIN, KDIGO, and RIFLE Definitions to Diagnose Acute Kidney Injury and Predict the Outcomes after Cardiac Surgery in a South Asian Cohort. Cardiorenal Medicine. 2022;12(1):29–38. doi:10.1159/000523828

39. Version EndNote 21. Clarivate; 2013.

40. Sterne JAC, Savović J, Page MJ, et al. RoB 2: a revised tool for assessing risk of bias in randomised trials. BMJ. 2019;366:l4898. doi:10.1136/bmj.l4898

41. Aromataris E MZE. JBI, ed. JBI Manual for Evidence Synthesis. 2020. https://synthesismanual.jbi.global. 10.46658/JBIMES-20-01

42. IntHout J, Ioannidis JPA, Borm GF. The Hartung-Knapp-Sidik-Jonkman method for random effects meta-analysis is straightforward and considerably outperforms the standard DerSimonian-Laird method. BMC Medical Research Methodology. 2014/02/18 2014;14(1):25. doi:10.1186/1471-2288-14-25

43. DerSimonian R, Laird N. Meta-analysis in clinical trials revisited. Contemporary clinical trials. 2015;45:139–145.

44. Sterne JA, Egger M. Funnel plots for detecting bias in meta-analysis: guidelines on choice of axis. J Clin Epidemiol. Oct 2001;54(10):1046–55. doi:10.1016/s0895-4356(01)00377-8

45. Egger M, Smith GD, Schneider M, Minder C. Bias in meta-analysis detected by a simple, graphical test. BMJ. 1997;315(7109):629–634. doi:10.1136/bmj.315.7109.629

46. Stata Statistical Software: Release 17 2021.

47. Schünemann HJ, Higgins JP, Vist GE, et al. Completing ‘Summary of findings’ tables and grading the certainty of the evidence. Cochrane Handbook for systematic reviews of interventions. 2019:375–402.

48. Page MJ, McKenzie JE, Bossuyt PM, et al. The PRISMA 2020 statement: An updated guideline for reporting systematic reviews. PLOS Medicine. 2021;18(3):e1003583. doi:10.1371/journal.pmed.1003583

49. Markewitz A, Hammer C, Pfeiffer M, et al. Reduction of cyclosporine-induced nephrotoxicity by cilastatin following clinical heart transplantation. Transplantation. 1994/03// 1994;57(6):865–870. doi:10.1097/00007890-199403270-00017

50. Hakeam HA, AlAnazi L, Mansour R, AlFudail S, AlMarzouq F. Does nephrotoxicity develop less frequently when vancomycin is combined with imipenem-cilastatin than with meropenem? A comparative study. Infectious Diseases. 2019/08/03 2019;51(8):578–584. doi:10.1080/23744235.2019.1619934

51. Shayan M, Elyasi S. Cilastatin as a protective agent against drug-induced nephrotoxicity: a literature review. Expert Opinion on Drug Safety. 2020/08/02 2020;19(8):999–1010. doi:10.1080/14740338.2020.1796967

52. Humanes B, Camaño S, Lara JM, et al. Cisplatin-induced renal inflammation is ameliorated by cilastatin nephroprotection. Nephrol Dial Transplant. 2017;32(10):1645–1655. doi:10.1093/ndt/gfx005

