## Supplementary material for "Nephroprotective effects of cilastatin in people at risk of acute kidney injury: A systematic review and meta-analysis": Figures

**Table S1 Literature search strategy applied to electronic bibliographic databases**

| **SN** | **Databases** |  |
| --- | --- | --- |
|  | **Database: Ovid MEDLINE(R) ALL <1946 to November 21, 2023> Search Strategy** | **Results** |
| 1 | exp Acute Kidney Injury/ | 56,927 |
| 2 | ("acute kidney injuries" or "kidney injur*or renal injur*" or "renal insufficienc*" or "kidney insufficienc*" or "kidney failure*").tw,kf. | 38,245 |
| 3 | exp Kidney Failure, Chronic/ | 101,630 |
| 4 | ("end-stage kidney disease*" or "end stage kidney disease*" or "chronic kidney failure" or "end-stage renal disease*" or "end stage renal disease*" or "end-stage renal failure" or "end stage renal failure" or "chronic renal failure" or "kidney dysfunction" or esrd).tw,kf. | 83,827 |
| 5 | (nephroprotection or nephroprotective* or prevention* or protection* or nephrotoxicity or "preventive measure*" or "all cause mortality" or mortality or death or "cardiovascular event*" or "kidney function*" or "kidney dysfunction" or creatinine or "cystatin c" or "glomerular filtration rate" or "urine output" or "allograft function" or proteinuria or albuminuria or NGAL or KIM-1 or Il-18 or L-FABP or IGFBP7 or TIMP-2).tw,kf. | 2,953,944 |
| 6 | 1 or 2 or 3 or 4 or 5 | 3,084,479 |
| 7 | exp Cilastatin/ or exp Cilastatin, Imipenem Drug Combination/ | 1,055 |
| 8 | (141a6amn38 or 5428wxz74m or 81129-83-1 or "mono-na salt" or "82009-34-5 cilastatin monosodium salt" or "cilastatin sodium" or "mk 0791" or mk 791 or "monosodium salt, cilastatin" or "salt, cilastatin monosodium" or "sodium, cilastatin").tw,kf. | 209 |
| 9 | 7 or 8 | 1,103 |
| 10 | 6 and 9 | 194 |
| 11 | animals/ not human/ | 5,139,807 |
| 12 | 10 not 11 | 161 |
|  | **Database: Embase <1974 to 2023 November 21> search strategy** |  |
| 1 | exp acute kidney failure/ | 125,050 |
| 2 | ("acute kidney injuries" or "kidney injur*or renal injur*" or "renal insufficienc*" or "kidney insufficienc*" or "kidney failure*").tw,kf. | 55,319 |
| 3 | exp chronic kidney failure/ | 155,047 |
| 4 | ("end-stage kidney disease*" or "end stage kidney disease*" or "chronic kidney failure" or "end-stage renal disease*" or "end stage renal disease*" or "end-stage renal failure" or "end stage renal failure" or "chronic renal failure" or "kidney dysfunction" or esrd).tw,kf. | 130,552 |
| 5 | (nephroprotection or nephroprotective* or prevention* or protection* or nephrotoxicity or "preventive measure*" or "all cause mortality" or mortality or death or "cardiovascular event*" or "kidney function*" or "kidney dysfunction" or creatinine or "cystatin c" or "glomerular filtration rate" or "urine output" or "allograft function" or proteinuria or albuminuria or NGAL or KIM-1 or Il-18 or L-FABP or IGFBP7 or TIMP-2).tw,kf. | 4,076,741 |
| 6 | exp cilastatin/ or exp cilastatin, imipenem drug combination/ | 8,053 |
| 7 | (141a6amn38 or 5428wxz74m or 81129-83-1 or "mono-na salt" or "82009-34-5 cilastatin monosodium salt" or "cilastatin sodium" or "mk 0791" or mk 791 or "monosodium salt, cilastatin" or "salt, cilastatin monosodium" or "sodium, cilastatin").tw,kf. | 457 |
| 8 | 1 or 2 or 3 or 4 or 5 | 4,278,218 |
| 9 | 6 or 7 | 8,075 |
| 10 | 8 and 9 | 1,864 |
| 11 | animals/ not human/ | 1,043,676 |
| 12 | 10 not 11 | 1,861 |
| 13 | limit 12 to (full text and abstracts and human) | 324 |
|  | **Database: Cochrane_search_2023-11-21**  **Search strategy** |  |
| 1 | [mh "acute kidney injury"] | 2056 |
| 2 | ("acute kidney injuries" or "kidney injury or renal injury" or "renal insufficiency" or "kidney insufficiency" or "kidney failure"):ti,ab,kw | 21152 |
| 3 | [mh “Kidney Failure, Chronic”] 5555 |  |
| 4 | (("end-stage kidney disease" or "end stage kidney disease" or "chronic kidney failure" or "end-stage renal disease" or "end stage renal disease" or "end-stage renal failure" or "end stage renal failure" or "chronic renal failure" or "kidney dysfunction")):ti,ab,kw | 13078 |
| 5 | ((nephroprotection or nephroprotective or prevention or protection or nephrotoxicity or "preventive measure" or "all cause mortality" or mortality or death or "cardiovascular event" or "kidney function" or "kidney dysfunction" or creatinine or "cystatin c" or "glomerular filtration rate" or "urine output" or "allograft function" or proteinuria or albuminuria or NGAL or KIM-1 or Il-18 or L-FABP or IGFBP7 or TIMP-2)):ti,ab,kw | 396102 |
| 6 | [mh Cilastatin] or [mh “Cilastatin, Imipenem Drug Combination”] | 229 |
| 7 | ("141a6amn38" or "5428wxz74m" or "81129-83-1" or "mono-na salt" or "82009-34-5 cilastatin monosodium salt" or "cilastatin sodium" or "mk 0791" or "mk 791" or "monosodium salt, cilastatin" or "salt, cilastatin monosodium" or "sodium, cilastatin"):ti,ab,kw | 37 |
| 8 | #1 or #2 or #3 or #4 or #5 | 406304 |
| 9 | #6 or #7 | 250 |
| 10 | #8 AND #9 | 67 |
|  | **Database: Web of Science (Nov 22, 2023)**  **Search Strategy** |  |
| 1 | ("acute kidney injuries" or "kidney injur*"or "renal injur*" or "renal insufficienc*" or "kidney insufficienc*" or "kidney failure*"). (Topic) | 101950 |
| 2 | TS=((“end-stage kidney disease*” or “end stage kidney disease*” or “chronic kidney failure” or “end-stage renal disease*” or “end stage renal disease*” or “end-stage renal failure” or “end stage renal failure” or “chronic renal failure” or “kidney dysfunction” or esrd)) | 86380 |
| 3 | TS=((nephroprotection or nephroprotective* or prevention* or protection* or nephrotoxicity or “preventive measure*” or “all cause mortality” or mortality or death or “cardiovascular event*” or “kidney function*” or “kidney dysfunction” or creatinine or “cystatin c” or “glomerular filtration rate” or “urine output” or “allograft function” or proteinuria or albuminuria or NGAL or KIM-1 or Il-18 or L-FABP or IGFBP7 or TIMP-2)) | 4007882 |
| 4 | #1 OR #2 OR #3 | 4094231 |
| 5 | TS=((Cilastatin or "Cilastatin, imipenem drug combination") or (141a6amn38 or 5428wxz74m or 81129-83-1 or “mono-na salt” or 82009-34-5 “Cilastatin monosodium salt” or “Cilastatin sodium” or mk 0791 or mk 791 or “monosodium salt, Cilastatin” or “salt, Cilastatin monosodium” or “sodium, Cilastatin”)) | 1802 |
| 6 | #4 AND #5 | 463 |

**List of studies included in systematic review and meta-analysis**

1.	Baghaie A BM, Abobo C, et al.,. The effect of imipenem/

cilastatin on acute cyclosporine nephrotoxicity in heart/lung

transplant patients. 1995:23:A241.

2.	Carmellini M, Frosini F, Filipponi F, Boggi U, Mosca F. Effect of cilastatin on cyclosporine-induced acute nephrotoxicity in kidney transplant recipients. Transplantation. 1997;64(1):164-6.

3.	Carmellini M, Matteucci E, Boggi U, Cecconi S, Giampietro O, Mosca F. Imipenem/cilastatin reduces cyclosporin-induced tubular damage in kidney transplant recipients. Transplant Proc. 1998;30(5):2034-5.

4.	Gruss E, Tomas JF, Bernis C, Rodriguez F, Traver JA, FernandezRanada JM. Nephroprotective effect of cilastatin in allogeneic bone marrow transplantation. Results from a retrospective analysis. Bone Marrow Transplant. 1996;18(4):761-5.

5.	Hakeam HA, AlAnazi L, Mansour R, AlFudail S, AlMarzouq F. Does nephrotoxicity develop less frequently when vancomycin is combined with imipenem-cilastatin than with meropenem? A comparative study. Infectious Diseases. 2019;51(8):578-84.

6.	Hornik CP, Herring AH, Benjamin DK, Jr., Capparelli EV, Kearns GL, van den Anker J, et al. Adverse events associated with meropenem versus imipenem/cilastatin therapy in a large retrospective cohort of hospitalized infants. Pediatric Infectious Disease Journal. 2014;32(7):748-53.

7.	Markewitz A, Hammer C, Pfeiffer M, Zahn S, Drechsel J, Reichenspurner H, Reichart B. Reduction of cyclosporine-induced nephrotoxicity by cilastatin following clinical heart transplantation. Transplantation. 1994;57(6):865-70.

8.	Schmitt DV, Leitner E, Welte T, Lode H. Piperacillin/tazobactam vs imipenem/cilastatin in the treatment of nosocomial pneumonia - a double blind prospective multicentre study. Infection. 2006;34(3):127-34.

9.	Zaballos M, Power M, Canal-Alonso MI, González-Nicolás MA, Vasquez-Jimenez W, Lozano-Lominchar P, et al. Effect of Cilastatin on Cisplatin-Induced Nephrotoxicity in Patients Undergoing Hyperthermic Intraperitoneal Chemotherapy. Int J Mol Sci. 2021;22(3):17.

10.	Zanetti G, Bally F, Greub G, Garbino J, Kinge T, Lew D, et al. Cefepime versus imipenem-cilastatin for treatment of nosocomial pneumonia in intensive care unit patients: A multicenter, evaluator-blind, prospective, randomized study. Antimicrob Agents Chemother. 2003;47(11):3442-7.

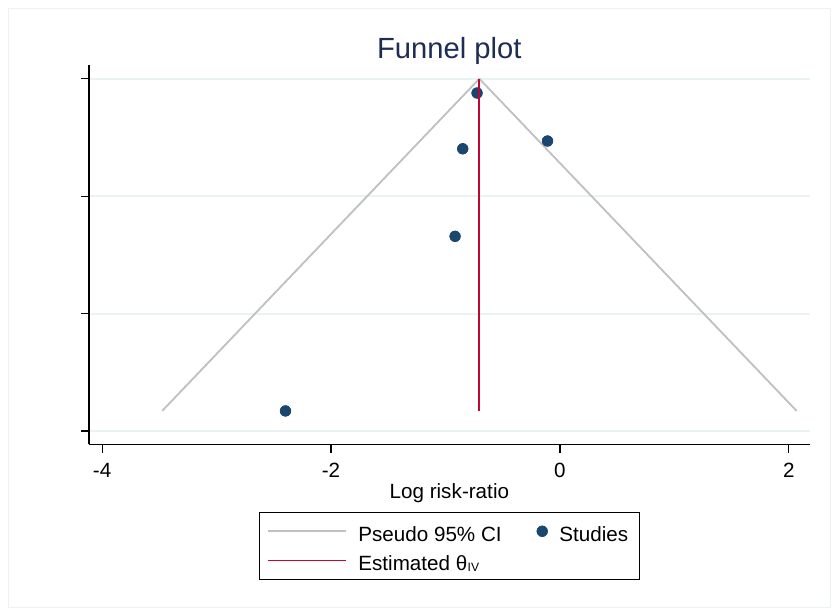

**A. Funnel Plot for AKI**

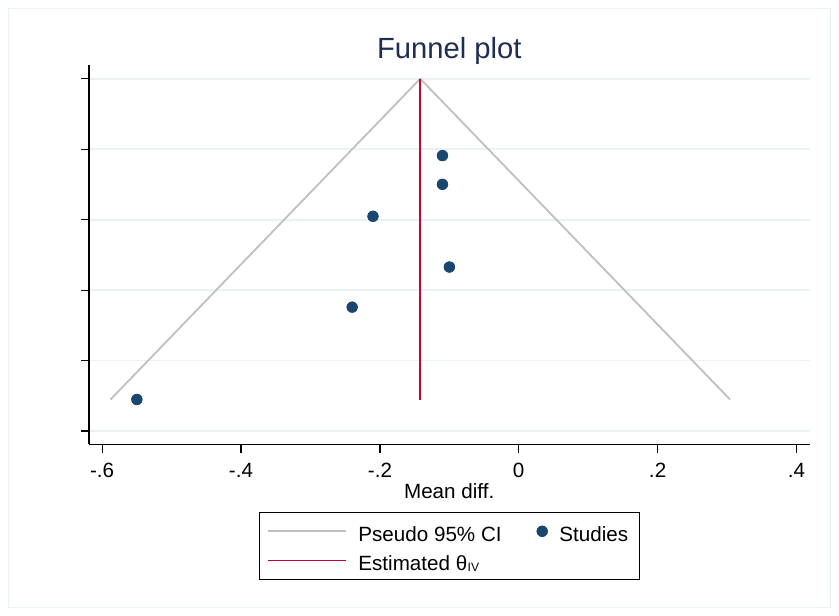

**B. Funnel Plot for Serum Creatinine**

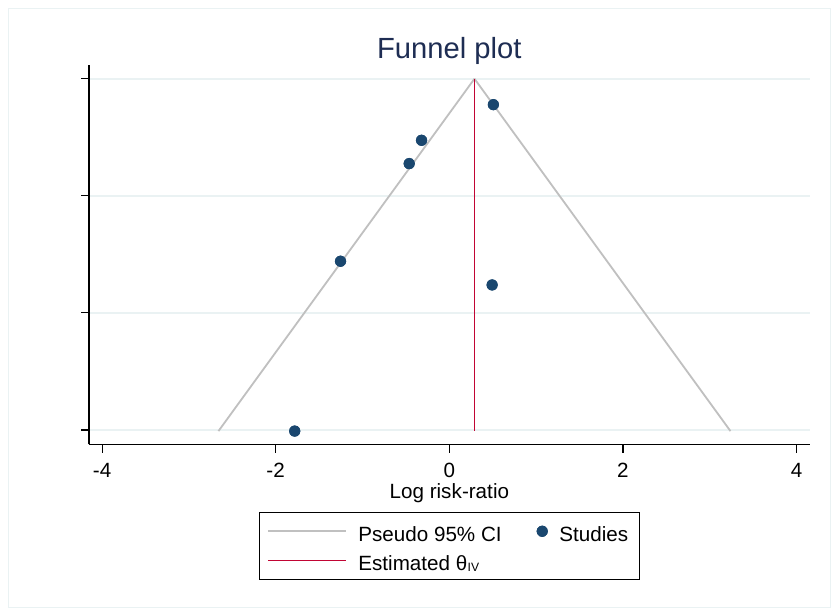

**C. Funnel Plot for All-cause Mortality**

**Supplementary Figure S1 Funnel plots for small study effects suggesting publication bias.**
